# DiGAS: Differential gene allele spectrum as descriptor in genetic studies

**DOI:** 10.1101/2023.10.16.23297102

**Authors:** Antonino Aparo, Vincenzo Bonnici, Simone Avesani, Luciano Cascione, Rosalba Giugno

**Affiliations:** University of Verona, Strada le Grazie, 15, Verona, 37134, Italy; University of Parma, Parco Area delle Scienze, 53/A, Parma, 43124, Italy; Institute of Oncology Research (IOR), Via Francesco Chiesa 5, Bellinzona, 6500, Switzerland

**Keywords:** genomic variations, alzheimer’s disease, classification, gene allele

## Abstract

Diagnosing subjects in complex genetic diseases is a very challenging task. Computational methodologies exploit information at genotype level by taking into account single nucleotide polymorphisms (SNP). They leverage the result of genome-wide association studies analysis to assign a statistical significance to each SNP. Recent methodologies extend such an approach by aggregating SNP significance at genetic level in order to identify genes that are related to the condition under study. However, such methodologies still suffer from the initial single-SNP analysis. Here, we present DiGAS, a tool for diagnosing genetic conditions by computing significance, by means of SNP information, but directly at the gene level. Such an approach is based on a generalized notion of allele spectrum, which evaluates the complete genetic alterations of the SNP set composing a gene at population level. Statistical significance of a gene is then evaluated by means of a differential analysis between the healthy and ill portions of the population. Tests, performed on well-established data sets regarding Alzheimer’s disease, show that DiGAS outperforms the state-of-the-art in distinguishing between ill and healthy subjects.

**Highlights:**

- We introduce a new generalized version of allele frequency spectrum.
- We propose a methodology, called DiGAS, based on the new defined genomic information and independent from GWAS analysis that out-performs existing methods in distinguish healthy/ill subjects with a speed up of 5x.
- On a reference Alzheimer’s disease genomic datasets, ADNI, DiGAS reaches F1 score up to 0.92.
- DiGAS methodology manages any type of genomic features, such as genes, exons, upstream/downstream regions.

## 1. Introduction

Human beings share more than 99 percent of their DNA sequence however, that small percentage of variation in DNA can have significant implications for human health. These variations can manifest as single nucleotide polymorphisms (SNPs), insertions, deletions, or larger rearrangements of DNA sequences, occurring both within and outside of genes. Single nucleotide polymorphisms (SNPs) are the most abundant type of genetic variation in the human genome, occurring approximately every 300 base pairs [1]. The primary focus on SNPs in genetic analysis is justified by their abundance, wide genomic coverage, heritability, functional impact, relevance in population studies, and clinical applications. SNPs, characterized by a single nucleotide substitution, follow Mendelian inheritance patterns and contribute to the heritability of diseases and traits [2]. By studying and analyzing the presence of one or more SNPs whether they occur within genes (intragenic) or in non-coding regions (intergenic), researchers gain insights into the underlying mechanisms of diseases. This understanding helps improve the assessment of disease risk, develop targeted therapies, and advance personalized medicine approaches [3, 4]. For instance, a specific single nucleotide polymorphism in the APOE gene has been found to influence the development of Alzheimer’s disease [5, 6], the deletion within the chemokine-receptor gene CCR5 provides resistance to HIV and AIDS [7]. Variations in genes related to immune responses can impact an individual’s susceptibility to autoimmune disorders or infectious diseases. [3]. Additionally, the identification of rare DNA variations has enabled the development of targeted therapies for cystic fibrosis [4].

GWAS (genome-wide association study) is a well-established methodology for associating genetic variants to disease risk in population genetics studies [8, 9]. This method identifies common variations that are present in the DNA sequences of individuals affected by a specific condition, under the hypothesis that common variants are present for the entire population. GWAS testing of millions of variants is often constrained by multiple hypothesis testing [10], as the analysis of a large number of hypotheses increases the likelihood of obtaining false-positive results.

Individual SNPs identified by GWAS platforms often show only modest effects. One reason is that the true causal SNP is rarely recognized, but there are SNPs that are in linkage disequilibrium (LD) with the causal SNP. In this case, when individual SNP analysis is used, the LD SNPs with the causal SNP will each show only moderate effects because each LD SNP acts as an imperfect surrogate for the causal SNP. Therefore, it might be advantageous to consider the joint effect of multiple SNPs in the analysis, because it is likely that many of these markers are in LD with the causal SNP and could capture the true effect more effectively than individual SNP analysis. Hundreds of studies have demonstrated that genes and their proteins often co-operate and interact together in functional pathways [11, 12]. Genes and SNPs could often predispose to disease through their reciprocal interaction in a specific biological pathway. Such a behaviour may produce a missing of these associations when a single-marker GWAS is used because of the relatively modest individual evidence of each gene/variant. Moreover, working at gene or pathway level reduces the number of possible tests, improves the statistical power, and might identify novel loci without increasing sample sizes or collecting new data. In addition, the probability that the result is true positive increases when combining supporting biological evidence with statistical significance. This means that it is advisable to restrict the analysis to candidate genomic regions (e.g. promoter regions, tissue-specific genes) or to prioritize candidate genes (e.g. having significant roles in specific pathways). In this perspective, SKAT [13] tests each SNP sets using a logistic kernel-machine regression framework to model the joint effect of the SNPs in the SNP sets. A SNP set can be any genomic region defined by users. SNPs are grouped according to their location in genomic features such as genes or haplotype blocks. The goal of SNP set analysis is to test the global null hypothesis of whether any of the SNPs are related to the outcome while adjusting for the additional covariates.

Similarly, other SNP sets analyses require the computation of gene-level p-values or gene scores. In [14], the SNP with the smallest p-value is used as representative of the entire gene. On the contrary, an empirical p-value for a SNP set is determined by recomputing the p-values of individual SNPs using a permuted dataset. The SNP set’s p-value is then calculated as the number of times where the average p-value of the observed SNPs is lower than the p-values obtained from the permuted data [15, 16, 17].

A similar empirical gene’s p-value is also computed in [18] but a multi-variate normal distribution is used to correct for linkage disequilibrium (LD) between SNPs. Alternatively, a null chi-square distribution is applied to capture LD between SNPs in a gene [19]. However, all these methods inherit from GWAS all the issues of assigning a significance to each SNP in a single-SNP analysing before grouping SNPs into sets. Table 1 summarizes the characteristics of the above approaches along with their limitations.

**Table 1:** A summary of the most commonly used SNP sets methods and limitations.

| Methods | Description | Limitations |
| --- | --- | --- |
| minSNP | Computes a gene score based on the smallest SNP p-value observed within the gene in a GWAS. | Biases may occur as longer genes tend to have lower gene scores. |
| permSNP | Involves permuting case-control labels in genotype data, recalculating SNP p-values, and computing empirical gene p-values using the observed and permuted data. | Computationally expensive for genome-wide data sets; gene score precision depends on the number of permutations. |
| VEGAS | takes into account the observed correlation between SNPs (LD) and simulates a specified number of statistics from which the resulting p-value is calculated. | Precision of gene scores depends on the number of simulations; computationally inefficient due to simulations. |
| Pegasus | Pegasus leverages pathway-based information to prioritize weak signals in GWAS. | Performance heavily influenced by the quality and the relevance of pathway databases. |
| SKAT | Employs mixed-model regression, considering covariates and genotypes for variants in a gene set to assess disease association. | May have limited power for small sample sizes and rare variant detection.<br>Assume linear relationships between SNPs and the phenotype. |

In this context, we introduce DiGAS, a tool that implements an innovative computational model for identifying genomic elements, ranging from individual exons to entire genomic regions may be associated with a given phenotype condition, such as a disease, and considered potential causal factors. The analysis involves the utilization of a novel genomic information descriptor termed the “generalized allele spectrum.” This descriptor is built upon the allele frequency spectrum, which captures allele frequencies within a defined group of loci (specifically, SNPs). The allele spectrum combines the frequency of single alleles into a unique vector of allele frequencies. In contrast to allele spectrum, the novel descriptor takes into account the complete set of SNPs of a region at once. This allows it to compute frequency at the region level rather than the SNP level. We define the the Differential Generalized Allele Spectrum to capture the differences in frequency allele spectra between healthy and ill sets (control and case respectively). The proposed methodology i) recognizes genetic regions that are important for a given pathology, and ii) builds a set of features for supervised classification purposes.

DiGAS represents a significant advancement over the state of the art, offering distinct advantages compared to both methods that aggregate SNPs individually and regression-based methods, such as SKAT (Sequence Kernel Association Test). Unlike methods that aggregate SNPs individually, DiGAS comprehensively analyzes the entire set of SNPs within genomic regions simultaneously. This approach allows DiGAS to capture potential joint effects of multiple SNPs, providing increased statistical power to identify genomic elements associated with the phenotype. In contrast, individual SNP aggregation methods may overlook such joint effects, potentially leading to a loss of relevant genetic associations. Moreover, DiGAS introduces the generalized allele spectrum descriptor, capturing genetic variations at the region level, thereby overcoming limitations of SNP-level analyses commonly employed by methods that aggregate SNPs individually. The generalized allele spectrum descriptor enables a more comprehensive representation of genomic variations, enhancing the accuracy of genetic signal attribution to specific genomic regions. Additionally, DiGAS generates interpretable outputs by identifying sets of features based on allele frequency differences. This feature selection process facilitates a clearer understanding of the genetic elements associated with the phenotype. In contrast, regression-based methods, such as SKAT, may not provide such interpretable outputs, making it challenging to interpret the specific genetic contributions to the phenotype. Furthermore, DiGAS adopts a non-linear approach, allowing the detection of complex genetic effects associated with the phenotype. This is in contrast to regression-based methods like SKAT, which often assume linear relationships between SNPs and the phenotype. The non-linear approach of DiGAS allows for the identification of non-linear genetic associations that are common in complex diseases, potentially providing valuable insights into the underlying genetic mechanisms. In conclusion, DiGAS represents a significant advancement over the state of the art by offering a more comprehensive, interpretable, and non-linear approach to identify genomic elements associated with phenotypes. Its simultaneous analysis of SNPs within genomic regions, utilization of the generalized allele spectrum descriptor, and non-linear approach contribute to its effectiveness in capturing genetic variations and improving the understanding of genetic contributions to complex diseases.

We tested DiGAS in the case of Alzheimer (AD) [20, 21], a progressive disease, where dementia symptoms gradually worsen over a number of years. AD’s has no cure, and it represents a challenge at the forefront of biomedical research [22]. The exact cause of AD’s disease is not fully understood, but it is believed to be a complex interplay of genetic, environmental, and lifestyle factors. Genetic factors play a significant role in the development and progression of AD, with variations in certain genes increasing the risk of developing the condition. Single nucleotide polymorphisms (SNPs) are the most common type of genetic variation and are variations in a single nucleotide base pair in the DNA sequence. In AD disease, it has been observed that a specific SNP may be present and associated with the disease in one affected individual but may not be present or associated with the disease in another affected individual. This means that the presence or absence of a single specific SNP is not sufficient to determine the disease status or predict its occurrence. Instead, AD disease is influenced by the combined effect of multiple SNPs that can vary from one individual to another. Each individual may have a unique combination of genetic variations, including different SNPs, that contribute to their susceptibility or resilience to the disease[23, 24].

The combined effect of multiple SNPs is thought to interact with other genetic, environmental, and lifestyle factors, leading to the complex and heterogeneous nature of Alzheimer’s disease. These factors may include variations in other genes, epigenetic modifications, interactions with environmental toxins, lifestyle choices, and overall health status. Understanding this concept highlights the need to investigate not only individual SNPs but also their interactions and cumulative effects. By considering the collective influence of multiple SNPs, researchers can gain a better understanding of the genetic architecture underlying the disease and potentially identify more comprehensive sets of genetic markers associated with Alzheimer’s disease risk and progression.

We compare DiGAS with SKAT since it allows to work with genotype data and thus be tested on different genomic regions. Results show that DiGAS outperforms SKAT in distinguishing healthy from ill subjects by means of their genomic features. Moreover, DiGAS remarkable reduces computational timing requirements compared to SKAT.

In what follows, Section 2.1 introduces the main methodological aspects of the proposed approach. Section 2.2 describes the datasets used for the evaluation of the proposed model. Finally, the results in the form of a supervised classification problem, are reported in Section 3. The DiGAS’s source code is freely available at https://github.com/InfOmics/DiGAS.

## 2. Material and Methods

In this section, we present the proposed methodology, DIGAS, along with details about the data used for testing and the validation approach.

Section 2.1 provides a formal description of the DIGAS method. A summary of the basic notions involved is reported in Table 2. The methodology involves the computation of the generalized allele spectrum, which is a measure related to the presence of SNPs in genomic regions for each phenotype condition analyzed in the study. Significant regions are identified based on the fold change of the generalized allele spectrum and the calculation of p-values using permutation tests.

**Table 2:** A summary of the terminology and notation used in this article.

|  |  |
| --- | --- |
| $\mathbb{S} = \{s_1, s_2, \dots, s_n\}$ | a population of $n$ individuals |
| $\mathbb{C} = \{c_1, c_2, \dots, c_j\}$ | a set of $j$ phenotype categories |
| $\mathbb{S}_c$ | a subset of $\mathbb{S}$ containing individuals belonging to category $c$ |
| $\gamma : \mathbb{S} \rightarrow \mathbb{C}$ | labelling of the individuals category |
| $\mathbb{P} = \{p_1, p_2, \dots, p_m\}$ | the set of $m$ SNPs taken into account in the study |
| $loc : \mathbb{P} \mapsto \mathbb{N}^+$ | the genomic location of a SNP |
| $\psi : \mathbb{S} \times \mathbb{P} \mapsto \{0, 1\}$ | the state (genetic variation present or absent) of each SNP for each subject |
| $\mathbb{G} = \{g_1, g_2, \dots, g_l\}$ | the set of regions that are investigated in the study |
| $\rho(g \in \mathbb{G}) = \{p_1, p_2, \dots, p_k\}$ | the set of SNPs whose genomic location reside within the genomic location of the region $G$ |
| $\eta_c(g) \in [0, 1]$ | the generalized allele spectrum of $g$ with respect to phenotype category $c$ |
| $FC_{c_1, c_2}(g)$ | the fold change of the generalized allele spectrum of region $g$ with respect to two categories $c_1$ and $c_2$ |

Section 2.2 describes the data used in the study and the preprocessing procedures applied to the data.

Finally, Section 2.3 provides a description of the classification algorithms and evaluation metrics used to assess the performance of the proposed DIGAS method.

DIGAS is implemented in Python. The method takes as input the coordinates of the genomic regions to be analyzed and the genotyping data (SNPs information). The DIGAS software is available for both Windows and Unix systems at the following GitHub repository: *https://github.com/InfOmics/DiGAS*.

### 2.1. DIGAS

Individuals with different phenotypic states can be categorized based on their conditions. For example, when studying a specific disease, we typically classify individuals into two groups: healthy and sick. However, it is also possible to consider more than two categories while ensuring that each individual belongs exclusively to one category.

In our model, the population of *n* individuals, referred to as subjects, is represented by the set 𝕊 ={ *s*_1_, *s*_2_, …, *s*_*n*_}, where *s*_*i*_ represents the *i*-th individual. To categorize these individuals, we have a set of categories ℂ = {*c*_1_, *c*_2_, …, *c*_*j*_}. We define a function *γ* : 𝕊 −*>* ℂ to assign a category to each subject. A subset of 𝕊 containing only the individuals belonging to category *c* ∈ ℂ is denoted as 𝕊_*c*_.

For each individual, we examine the occurrence of genomic single nucleotide variations, known as single nucleotide polymorphisms (SNPs), in relation to a selected reference genome. We establish the function *loc* : ℙ → ℕ^+^ to determine the position of a SNP within the genome. We define ℙ = {*p*_1_, *p*_2_, …, *p*_*m*_} as the set of *m* SNPs that are being considered. It is important to note that in diploid genomes, where two alleles are present for each genomic locus, we do not differentiate between diploid variations at the same locus.

The function *ψ* : 𝕊 ×ℙ ↦{0, 1} indicates the absence or presence of a SNP for an individual.

For instance, given an individual *s*_*i*_ ∈ 𝕊 and a SNP *p*_*j*_ ∈ ℙ, *ψ*(*s*_*i*_, *p*_*j*_) is 0 if no SNP is observed at *loc*(*p*_*j*_) in the genome of the individual *s*_*i*_.

Experimental designs may necessitate the detection of SNPs throughout the entire genome or in specific regions such as genes, exons, or intergenic regions. The scope of SNP detection can be tailored based on the objectives of the study and the specific genomic regions of interest.

Consider the set of regions to investigate as 𝔾 = {*g*_1_, *g*_2_, …, *g*_*l*_}, where each *g*_*i*_ represents a contiguous region of nucleotides defined by start and end coordinates with respect to the reference genome. We denote the subset of SNPs residing in the region *g*_*i*_ of the reference genome as *ρ*(*g*_*i*_ ∈ 𝔾) = ℙ_*i*_ ⊆ℙ.

This subset ℙ_*i*_ consists of SNPs where the genomic location *loc*(*p*_*j*_) satisfies the condition *start*(*g*_*i*_) ≤ *loc*(*p*_*j*_) ≤ *end*(*g*_*i*_) for each SNP *p*_*j*_ ∈ ℙ_*i*_. In simpler terms, ℙ_*i*_ includes SNPs located within the boundaries of the region *g*_*i*_ in the reference genome.

For a genomic region *g* belonging to the set 𝔾, the overall allele spectrum of *g* in relation to the specified phenotype category *c* represents the ratio between the total count of SNPs observed in the region across all individuals within that category and the maximum possible count of SNPs in that region for the same category. This can be defined as:

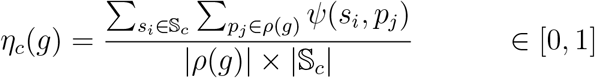

with *η*_*c*_(*g*) is in [0, 1], because *ψ*(*s*_*i*_, *p*_*j*_) can be 0 or 1 and the summation can not exceed |*ρ*(*g*) |×| 𝕊_*c*_|. The value is 1 when all subjects belonging to the given category present all SNPs in the considered region.

We aim to find genomic regions having statistically significant different values of allele spectrum among categories. For such propose, we define the fold change *FC* of a genomic region *g* with respect to two categories *c*_1_ and *c*_2_ as:

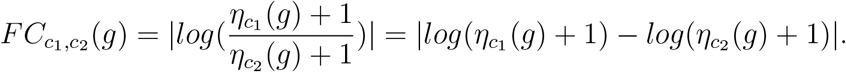

Our model computes the fold change of each region across each pair of phenotype categories. The selection of regions that are considered significant is obtained by calculating an empirical *p*-value using a permutation test [25]. To achieve this, we initiate the process by randomly permuting the original category assignments of the subjects. This results in the creation of 1000 different random labelings of subject categories, denoted as {*γ*_0_, *γ*_1_, …, *γ*_1000_}. To determine the significance of the observed fold change in the real data, we calculate the proportion of random labelings where the fold change is equal to or greater than the observed value. This proportion represents the p-value of the region. A lower p-value indicates that the observed fold change is less likely to occur by random chance alone, suggesting that the region may have a significant association with the categories being studied.

More precisely, we modify the original category assignment *γ* to a new function *γ*_*i*_, where the assignments in *γ*_*i*_ are a permutation of the assignments in *γ*. Thus, the total number of subjects assigned to each category, given two categories *c*_1_ and *c*_2_, is maintained from *γ* to *γ*_*i*_.

Let 𝕊*c*_1_ and 𝕊*c*_2_ be the subsets obtained according to the category assignments in *γ*. To obtain *γ*_*i*_, we iteratively modify *γ* for a total of 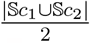 iterations. We refer to 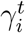 as the version of *γ*_*i*_ at iteration *t*, where 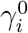 is an exact equal to *γ*. For each iteration *t >* 0, we select two subjects *s*_1_ and *s*_2_ such that 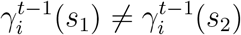. We create 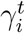 by swapping the assignments of *s*_1_ and *s*_2_, i.e., 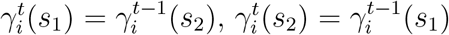, and 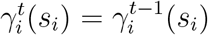 for *s*_*i*_ ∈ 𝕊 \ *s*_1_, *s*_2_.

The *p*-value of a region *g* is then determined by calculating the percentage of random labelings for which the fold change of the region equals or exceeds 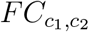. Regions that have a p-value less than 0.05 are considered relevant for discriminating between subjects who belong to category *c*_1_ from subjects who belong to category *c*_2_.

### 2.2. Test Dataset

The data used in this manuscript was obtained from The Alzheimer’s Disease Neuroimaging Initiative (ADNI) project (http://adni.loni.usc.edu). The ADNI researchers collect, validate, and utilize various types of data including MRI and PET images, genetics, cognitive tests, CSF (cerebrospinal fluid), and blood biomarkers to study and predict the disease. Our focus is on identifying regions of genomes which sets of SNPs collectively may contribute to the disease. Coordinates of the regions to take into account are provided by the GENCODE project^1^ (*v36lift37*). We considered the complete set of ADNI cohorts, which includes ADNI1, ADNI2/GO, and ADNI3. The individuals in these cohorts are classified into three categories: affected (AD), not affected (CN), and mild cognitive impairment (MCI). The MCI category encompasses individuals who exhibit symptoms similar to those of Alzheimer’s disease but do not exhibit a strong hallmark phenotype. In some cases, individuals with MCI may revert to normal conditions [26].

We filtered out all the individuals with no European ancestry. Statistics regarding the subjects extracted from ADNI are reported in Table 3. Quality control (QC) procedures were conducted on the data from each ADNI cohort using PLINK 1.9 format[27], which is a comprehensive toolset for whole-genome association analysis. These QC procedures involved filtering SNPs and subjects based on the following specific criteria. (i) Missing Data Filter (*geno >* 0.2): SNPs with a high proportion of missing data, where more than 20% of the data was missing, were excluded from the analysis. (ii)Individual Missingness Filter (*mind >* 0.1): SNPs were filtered based on individual missingness, where SNPs with more than 10% of individuals having missing genotype data were excluded.(iii) Minor Allele Frequency Filter (*MAF >* 0.05): SNPs with a minor allele frequency below 5% were removed. This filter helps to ensure that the analysis focuses on common genetic variations. (iv) Hardy-Weinberg Equilibrium Filter (*hwe >* 1e−06): SNPs showing significant deviations from the Hardy-Weinberg equilibrium were excluded. Hardy-Weinberg equilibrium represents the expected frequencies of genotypes in a population, and deviations from this equilibrium may indicate potential genotyping errors or other issues.

**Table 3:**
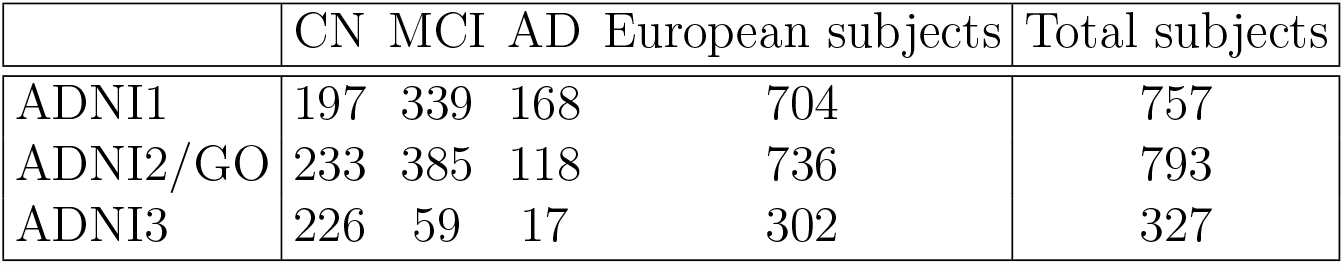
Number of European subjects (divided by categories) used as input for each ADNI cohort. Total number of subjects, independently from their ancestry, is also reported.

|  | CN | MCI | AD | European subjects | Total subjects |
| --- | --- | --- | --- | --- | --- |
| ADNI1 | 197 | 339 | 168 | 704 | 757 |
| ADNI2/GO | 233 | 385 | 118 | 736 | 793 |
| ADNI3 | 226 | 59 | 17 | 302 | 327 |

Table 4 provides information on the SNPs that were filtered out after applying these QC procedures. Regarding subjects, no individuals were filtered out based on QC measures. This means that all individuals in the ADNI cohorts were retained for further analysis after the QC procedures.

**Table 4:**
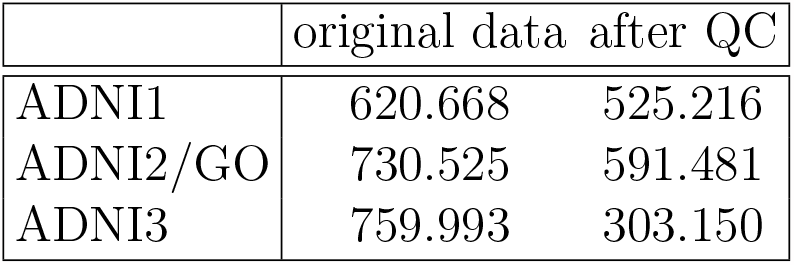
Total number of SNPs for each cohort and number of SNPs filtered by Quality Control (QC) procedures.

### 2.3. Evaluation methodology

We used a set of classification algorithms, such as linear discriminant analysis (LDA) [28], support-vector machine (SVM) [29] (linear and polynomial), decision tree [30] and k-nearest neighbors (k-NN) [31] in order to evaluate the ability of the proposed methodology in selecting regions that are useful for distinguishing subject’s categories. The goal of the classification is to build a model that, after a learning phase, correctly assigns a category to a given subject.

Given an input dataset, we applied a 2-fold cross-validation [32] which splits the original cohort into two subsets. One of the two subsets is used for training the classification model, and the other subset is used for validating the trained model. The split is done via a random selection of the subjects.

The selection ensures that the initial proportions among categories of the subjects are preserved.

More precisely, after the training phase, the resultant model is queried by using records belonging to the test set. A test set individual that is correctly recognized as belonging to a given category *C* by the model is considered a true positive (TP) for such a category. On the contrary, a false positive (FP) record is labelled as *C* by the model but in reality it does not belong to *C*. Similarly, true negative (TN) are records that are correctly classified as non-*C*, and false negative (FN) are records that are wrongly classified as not belonging to *C*.

Accuracy is defined as the fraction of records that are correctly classified with respect to the entire test set. The F1 score combines precision and recall statistics into a single metric via harmonic mean. Precision informs about the fraction of records that are correctly classified as belonging to *C* with respect to the total number of records that are classified as *C* by the model. Recall gives the fraction of records belonging to *C* that are correctly classified with respect to the total size of *C*.

All the given metrics are in the range of [0, 1] such that the higher the value, the better the performance of the given model is. Moreover, for binary classification, precision and recall are related to the given category that is taken into account. On the contrary, the value of accuracy is the same independently for the investigated category.

### 3. Results

We evaluated the efficacy of the DiGAS methodology in classifying Alzheimer’s disease subjects also with respect to SKAT [13].

Given an ADNI cohort (see Section 2.2 for details regarding the composition of the ADNI data set), we split the input data set using two different partition percentages, 90-10 and 70-30. This means that, if the ADNI1 cohort has 197*/*704 = 28% CN subjects, 48% MCI and 23% AD, such percentages are preserved in both the training and the validation sets. The partition 90-10 is intended to boost the efficacy of the approach at better performance, but it may incur over-fitting problems. For this reason, we decided to show here only the results on the partition 70-30. However, the evaluation performed by using 90-10 of the data set reflects the results obtained with the split at 70-30.

SNP sets are the features of our classification model. Thus, the goal is to recognize the SNP sets which make a distinction between the categories. To do so, we group SNPs by the following genomic regions:

- Exons: each exon is intended to be a single specific region, not linked to the exons of the same gene. Exon may belong to any type of gene, protein-coding or not.
- Protein-coding exons: namely exons that belong to genes which are known to code for proteins.
- Upstream exon regions: for each exon we extract 5k nucleotides that precede the exon. The exon is excluded from the extracted region.
- Exons+upstream: for each exon, we take into account the exon itself plus the upstream 5K nucleotides region.
- Genes: the complete genomic sequence of each gene, exons plus introns, is taken into account.
- Genes+upstream+downstream: we extract the upstream and the downstream, for 20Kb each, and the sequence of the gene itself. Such a setup equals the one used in [13].

The coordinates of such genomic elements are extracted from public databases described in Section 2.2. The belonging of a SNP to a given region is calculated via the *loc* function described in Section 2.1. All the experiments are performed over the GrCh38 version of the human genome. Since ADNI1 is originally defined over previous versions of the human genome, we used the tool UCSC LiftOver [33, 34] to convert such coordinates into coordinates over the GrCh38 genome.

We applied the methodology described in Section 2.1 for identifying significant regions. In particular, we applied a cut-off for the p-value (evaluated by means of the fold-change) of 0.05. In this process, the three categories, *CN, AD* and *MCI*, were evaluated separately. Then, we merged the regions that resulted significant for *AD* and *MCI* into a single set of regions. For this reason, in what follows, ill subjects are also referred to as the joint category *AD/MCI*.

For each cross-validation, the resultant performance metrics were calculated by running 1,000 iterations, and by computing the mean and the standard deviation of the results.

Figure 1 shows the accuracy values of DiGAS and SKAT on the ADNI1 cohort varying the genomic regions and the type of classifier. For each type of region, DiGAS always outperforms SKAT in particular using SVM classifiers. Independently from the type of genomic region, the classifier implemented by means of a decision tree shows the worst accuracy for both compared approaches. SKAT reaches a maximum of 0.78 when exons are used as the basis for training an SVM linear classifier, but in general SKAT accuracy is almost always below 0.75. On the contrary, DiGAS is able to break the barrier of 0.75 in multiple configurations. The best accuracy value of 0.94 is obtained when upstream exon regions are taken into account alone or in combination with exons and by using an SVM classifier.

**Figure 1:**
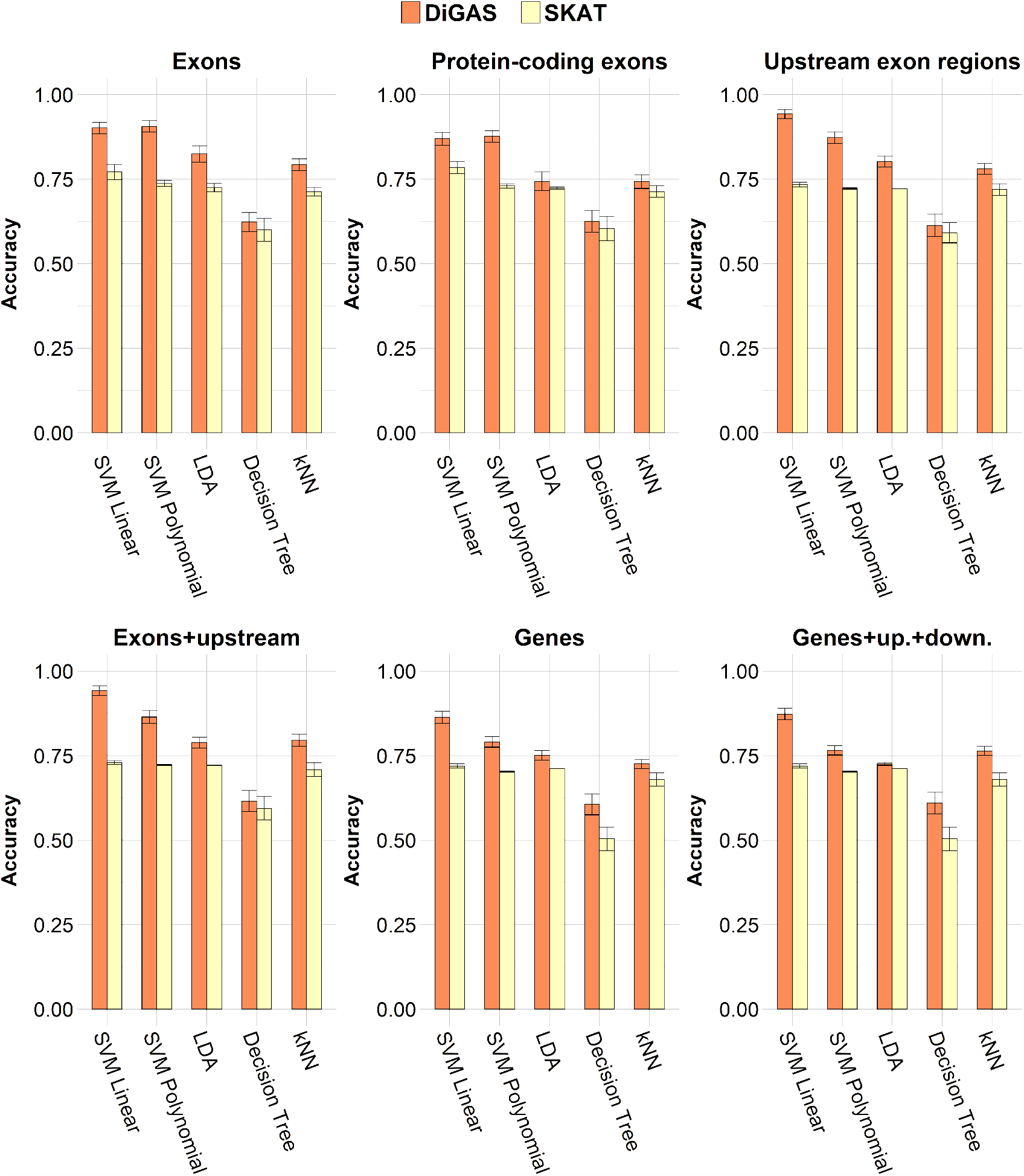
Accuracy metrics on ADNI1 by using 70% of the data as training set for each evaluated classification algorithm and each genomic region.

Similar results are shown for the F1 score for the ADNI1 cohort in Figure 2, with only one exception given by the kNN classifier on genes where SKAT outperforms DiGAS. SKAT reaches a maximum of 0.66 via exons, while DiGAS obtains up to a 0.92 of F1 score on both upstream exon regions and exon+upstream.

**Figure 2:**
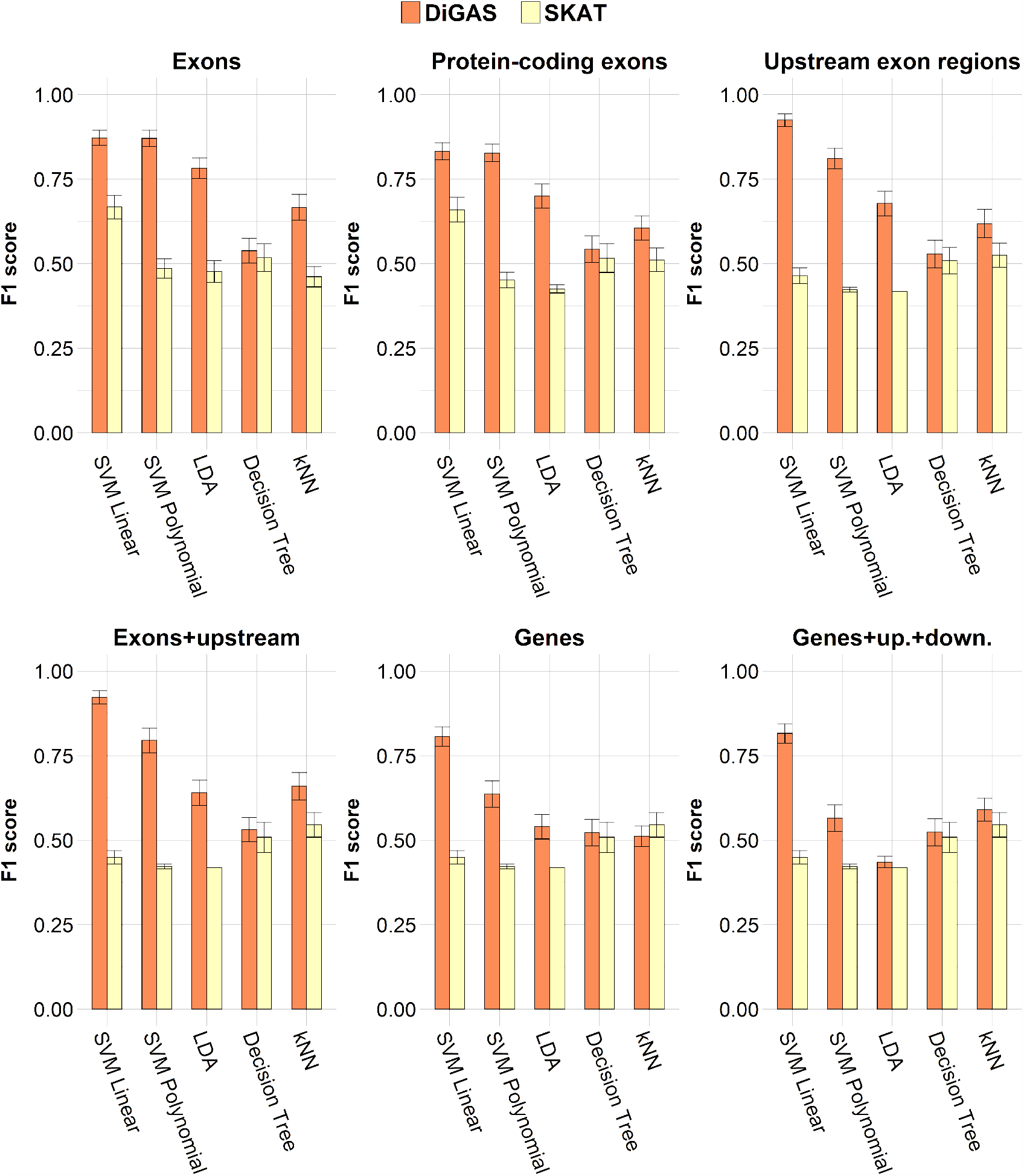
F1 score metrics on ADNI1 by using 70% of the data as training set for each evaluated classification algorithm and each genomic region.

Figures 3 and 4 report accuracy and F1 score values, respectively, on the ADNI2 cohort. These results reflect the performance obtained for the ADNI1 cohort. Maximum values of accuracy are 0.92 (exons+upstream) and 0.77 (exons) for DiGAS and SKAT, respectively. Maximum F1 scores are 0.90 (exons+upstream) and 0.70 (exons) for DiGAS and SKAT, respectively.

**Figure 3:**
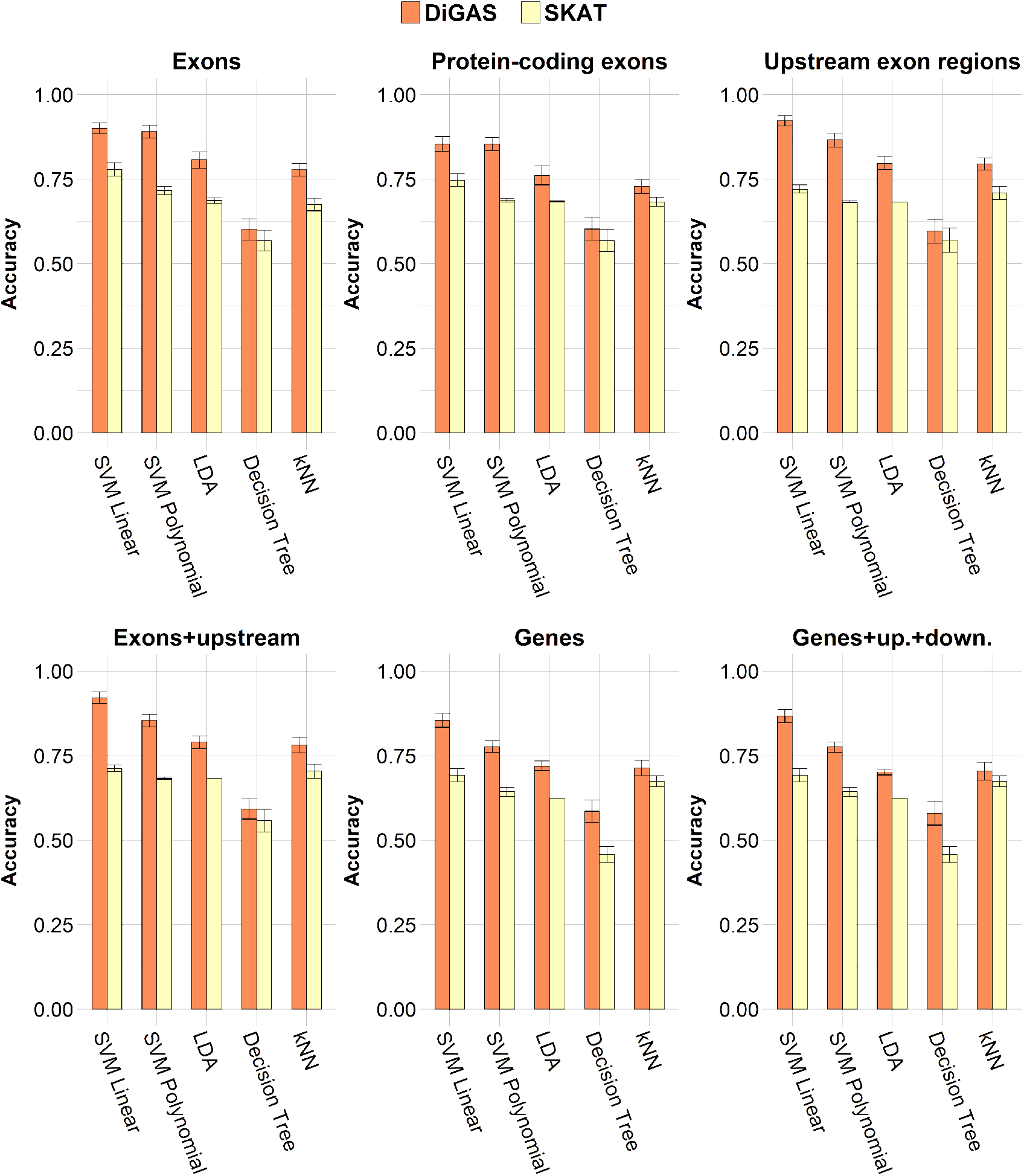
Accuracy metrics on ADNI2 by using 70% of the data as training set for each evaluated classification algorithm and each genomic region.

**Figure 4:**
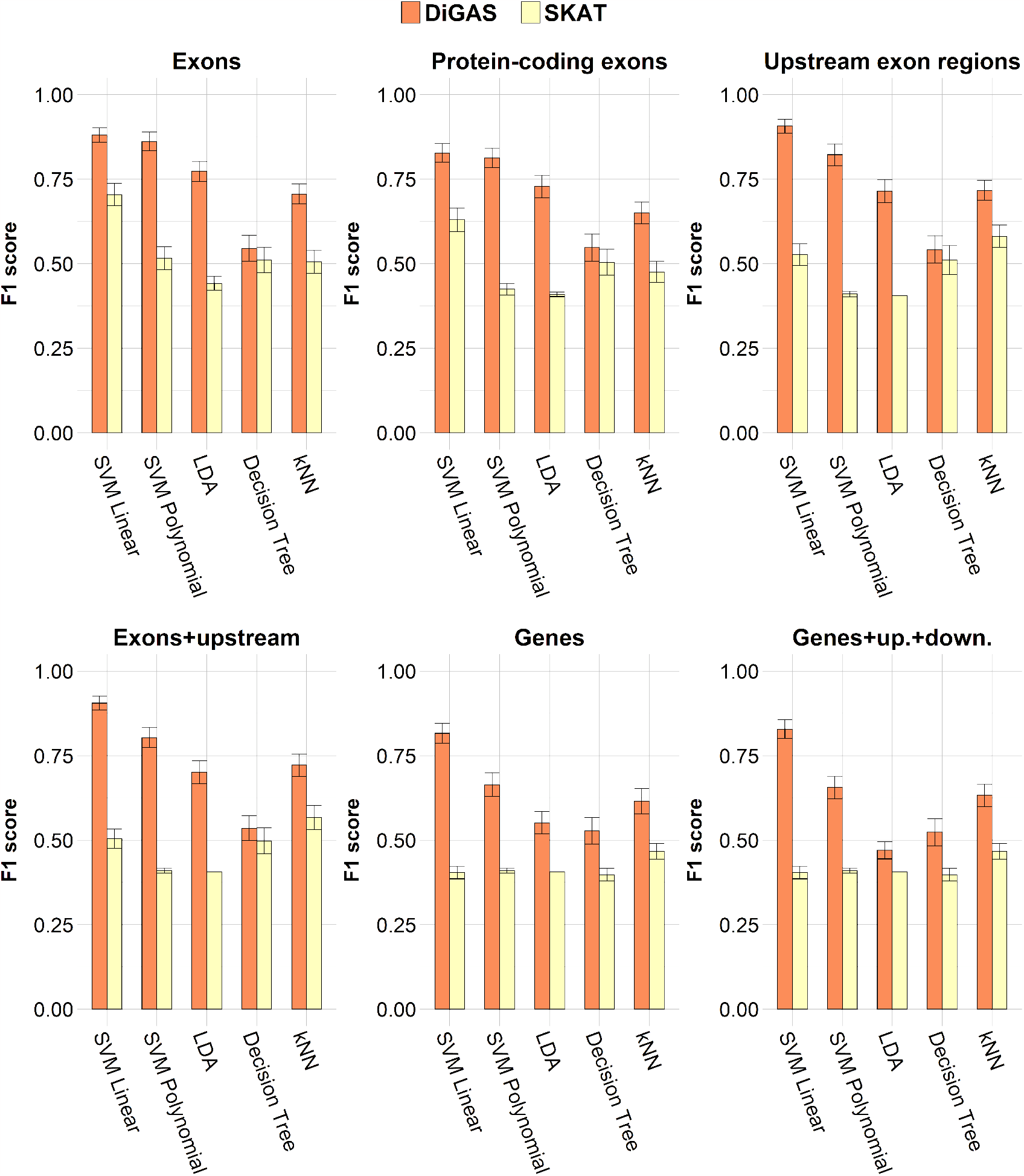
F1 score metrics on ADNI2 by using 70% of the data as training set for each evaluated classification algorithm and each genomic region.

Figures 5 and 6 show results obtained on the ADNI3 cohort. Accuracy values follow similar trends obtained by testing the methodologies on ADNI1 and ADNI2. On the contrary, DiGAS and SKAT reduce their performance on the F1 score. The difference with previous cohorts is due to the limited number of *AD/MCI* subjects that are in the data set. ADNI3 cohort is an ongoing project for which fewer ill subjects are yet reported. F1 scores for the *AD/MCI* group suffer such a lack of data that does not affect accuracy because such a measure takes into account both *CN* and *AD/MCI* groups. However, it has to be noticed that DiGAS is still able to reach an F1 score of 0.91 when exons regions are combined with the SVM linear classifier, and exons+upstream regions still produce a maximum value of 0.85 and 0.87 when SVM linear and kNN classifiers are employed. Moreover, DiGAS crosses the barrier of 0.70 in several configurations. On the contrary, SKAT reaches an F1 score greater the 0.70 only in three configurations, being the best one equal to 0.71 by combining exon regions with the SVM linear classifier. Thus, such results show that DiGAS, in contrast to SKAT, is more robust when a limited amount of information is available. In general, the SVM linear classifier is the best choice to work with the DiGAS methodology, but the kNN approach can be taken into account in the presence of data set with a category containing a limited number of subjects.

**Figure 5:**
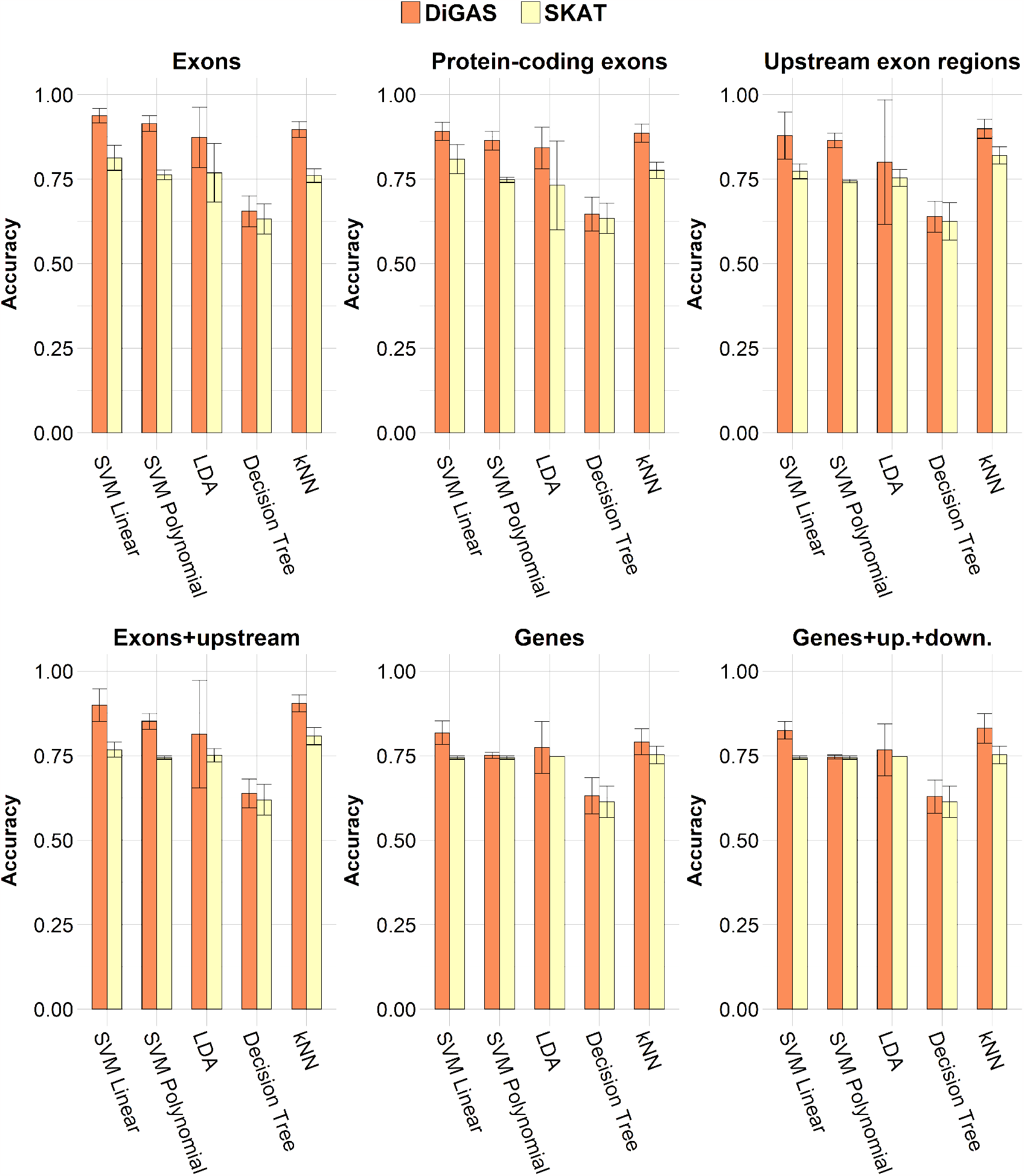
Accuracy metrics on ADNI3 by using 70% of the data as training set for each evaluated classification algorithm and each genomic region.

**Figure 6:**
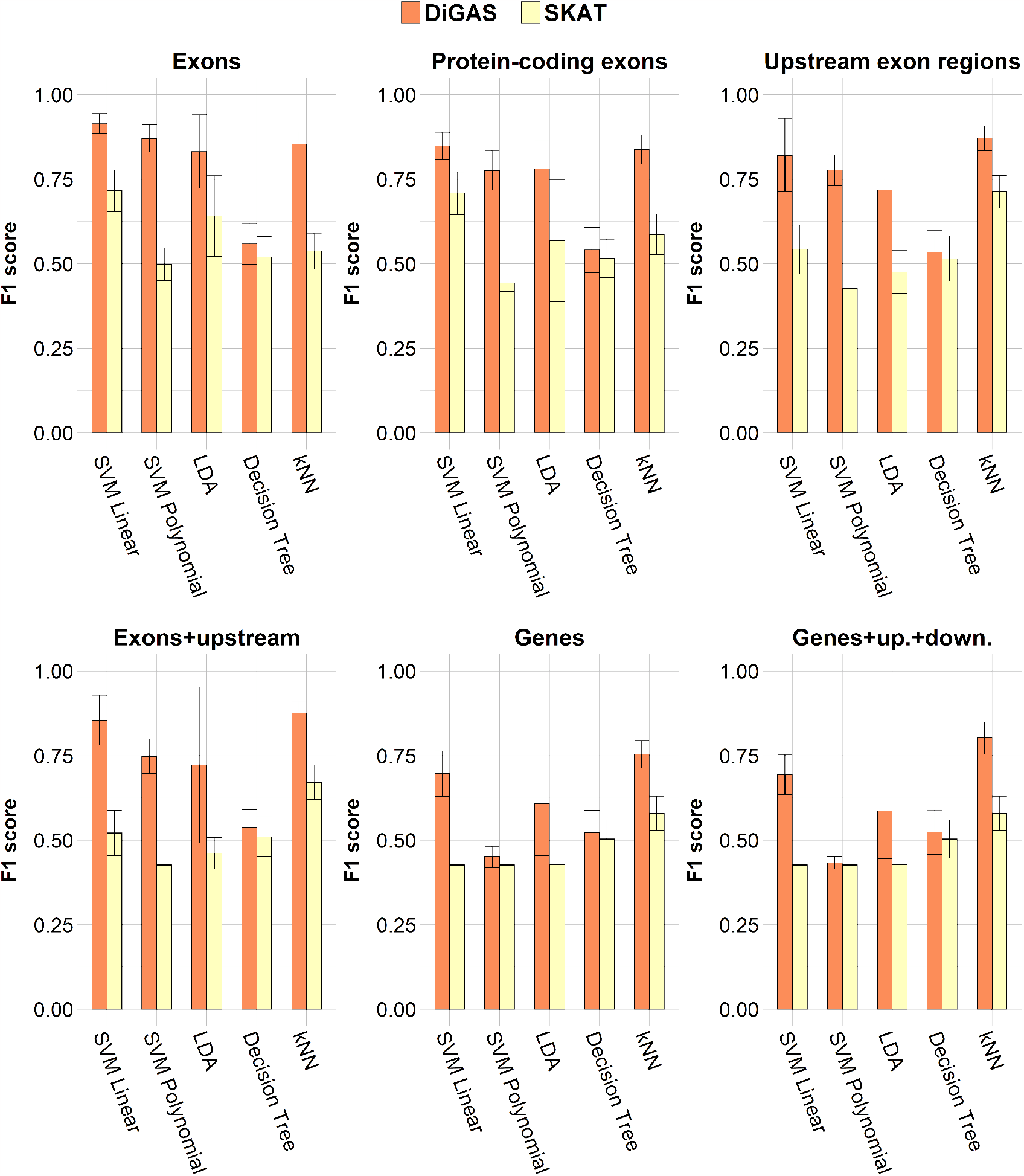
F1 score metrics on ADNI3 by using 70% of the data as training set for each evaluated classification algorithm and each genomic region.

Considering the required computing time resources, DiGAS is 5 orders of magnitude faster than SKAT.

In general, we note that exons, and in particular not only protein-coding exons, combined with upstream regions produce the best classification results. Alzheimer’s disease is a considered complex disease which involves many genes and, presumably, their regulatory elements [35, 36]. Such regulatory elements are often placed in upstream gene regions. However, our analysis shows that regulatory regions of genes are important as well as upstream exon regions. It is known that too much information may reduce classifiers performance, especially when such overabundant data does not relate with the recognition problem that is taken into account. The low performance on genes and their combination with upstream and downstream regions may reflect the importance of upstream exon regions, and thus inter- and intra-genic regulatory elements in Alzheimer’s disease, rather than the entire genetic sequence. In fact, upstream regions of exons alone produce comparable results when combined with exon sequences. Such upstream regions may overlap with exon regions, thus information contained in exons is taken into account in both cases. However, pure exon regions are outperformed by their combination with upstream regions.

## 4. Conclusions

In conclusion, we presented a methodology, DiGAS, for diagnosing complex genetic diseases, such as the Alzheimer’s disease, by means of phenotype data. Existing approaches are based on the results of GWAS analysis to assign a p-value to each SNP, then they aggregate SNP p-values at SNP set level. Differently from such approaches, DiGAS computes a SNP set p-value, according to the SNPs present in each set directly from genotype data. Tests, performed on well-established data sets regarding the Alzheimer’s disease, show that DiGAS outperforms the state-of-the-art method named SKAT in classification power and computational timing required.

## Data Availability

All data produced in the present study are available upon reasonable request to the authors

https://github.com/InfOmics/DiGAS

## Footnotes

1 https://www.gencodegenes.org

## Notes

### Competing Interest Statement

The authors have declared no competing interest.

### Funding Statement

This study did not receive any funding

